## Supplementary material for "Assessing the relationship between menstrual products and reproductive and urogenital tract infections (RUTIs): a systematic review evaluating the evidence and recommendations for future research": S1 Table & S2 Table

Supplementary Table 1. Full Literature Search Strategy for PubMed. Adapted for Entry into Web of Science

| **Outcome** | **Search String** |
| --- | --- |
| Bacterial Vaginosis | ("Menstrual Hygiene Products"[MeSH] OR "menstrual product*"[MeSH] OR "menstrual napkin*"[Title/Abstract] OR "period product*"[Title/Abstract] OR "feminine hygiene product*"[Title/Abstract] OR "feminine hygiene"[Title/Abstract] OR "feminine product*"[Title/Abstract] OR "feminine napkin*"[Title/Abstract] OR "sanitary pad*"[Title/Abstract] OR "menstrual cup"[Title/Abstract] OR "menses cup"[Title/Abstract] OR "menstruation cup"[Title/Abstract] OR "vaginal cup"[Title/Abstract] OR "menstrual cups"[Title/Abstract] OR "menses cups"[Title/Abstract] OR "menstruation cups"[Title/Abstract] OR "vaginal cups"[Title/Abstract] OR "reusable pad*"[Title/Abstract] OR ("menstrual" AND "cloths") OR ("menses" AND "cloths") OR ("menstruation" AND "cloths") OR ("vaginal" AND "cloths") OR ("menstruation"[Title/Abstract] AND "management"[Title/Abstract]) OR (("menstruation"[Title/Abstract] OR "menstrual"[Title/Abstract]) AND "product*"[Title/Abstract] AND "safety"[Title/Abstract]) OR "catamenia") AND ("Bacterial Vaginosis"[MeSH Terms] OR "bacterial vaginosis"[Title/Abstract] OR "BV"[Title/Abstract] OR "vaginal bacteriosis"[Title/Abstract] OR "Gardnerella"[Title/Abstract] OR "vaginal flora imbalance"[Title/Abstract] OR "vaginal dysbiosis"[Title/Abstract]) |
| Urinary Tract Infections | ("Menstrual Hygiene Products"[MeSH] OR "menstrual product*"[MeSH] OR "menstrual napkin*"[Title/Abstract] OR "period product*"[Title/Abstract] OR "feminine hygiene product*"[Title/Abstract] OR "feminine hygiene"[Title/Abstract] OR "feminine product*"[Title/Abstract] OR "feminine napkin*"[Title/Abstract] OR "sanitary pad*"[Title/Abstract] OR "menstrual cup"[Title/Abstract] OR "menses cup"[Title/Abstract] OR "menstruation cup"[Title/Abstract] OR "vaginal cup"[Title/Abstract] OR "menstrual cups"[Title/Abstract] OR "menses cups"[Title/Abstract] OR "menstruation cups"[Title/Abstract] OR "vaginal cups"[Title/Abstract] OR "reusable pad*"[Title/Abstract] OR ("menstrual" AND "cloths") OR ("menses" AND "cloths") OR ("menstruation" AND "cloths") OR ("vaginal" AND "cloths") OR ("menstruation"[Title/Abstract] AND "management"[Title/Abstract]) OR (("menstruation"[Title/Abstract] OR "menstrual"[Title/Abstract] OR "catamenia"[Title/Abstract]) AND "product*"[Title/Abstract] AND "safety"[Title/Abstract])) AND ("Urinary Tract Infections"[MeSH] OR "UTI"[Title/Abstract] OR "urogenital infections"[Title/Abstract] OR "urinary infection*"[Title/Abstract] OR "bladder infection*"[Title/Abstract]) |
| Sexually Transmitted Infections | ("Menstrual Hygiene Products"[MeSH] OR "menstrual product*"[MeSH] OR "menstrual napkin*"[Title/Abstract] OR "period product*"[Title/Abstract] OR "feminine hygiene product*"[Title/Abstract] OR "feminine hygiene"[Title/Abstract] OR "feminine product*"[Title/Abstract] OR "feminine napkin*"[Title/Abstract] OR "sanitary pad*"[Title/Abstract] OR "menstrual cup"[Title/Abstract] OR "menses cup"[Title/Abstract] OR "menstruation cup"[Title/Abstract] OR "vaginal cup"[Title/Abstract] OR "menstrual cups"[Title/Abstract] OR "menses cups"[Title/Abstract] OR "menstruation cups"[Title/Abstract] OR "vaginal cups"[Title/Abstract] OR "reusable pad*"[Title/Abstract] OR ("menstrual" AND "cloths") OR ("menses" AND "cloths") OR ("menstruation" AND "cloths") OR ("vaginal" AND "cloths") OR ("menstruation"[Title/Abstract] AND "management"[Title/Abstract]) OR (("menstruation"[Title/Abstract] OR "menstrual"[Title/Abstract]) AND "product*"[Title/Abstract] AND "safety"[Title/Abstract]) OR "catamenia") AND ("Sexually transmitted disease"[MeSH Terms] OR "Gonorrhea"[Title/Abstract] OR "Syphilis"[Title/Abstract] OR "Chlamydia"[Title/Abstract] OR "Sexually transmitted infections" [Title/Abstract]) |
| HIV | ("Menstrual Hygiene Products"[MeSH] OR "period product*"[Title/Abstract] OR "feminine hygiene product*"[Title/Abstract] OR "feminine hygiene"[Title/Abstract] OR "feminine product*"[Title/Abstract] OR "feminine napkin*"[Title/Abstract] OR "sanitary pad*"[Title/Abstract] OR "menstrual cup"[Title/Abstract] OR "vaginal cup"[Title/Abstract] OR "menstrual cups"[Title/Abstract] OR "menses cups"[Title/Abstract] OR "vaginal cups"[Title/Abstract] OR "reusable pad*"[Title/Abstract] OR ("menstrual"[All Fields] AND "cloths"[All Fields]) OR ("menses"[All Fields] AND "cloths"[All Fields]) OR ("menstruation"[All Fields] AND "cloths"[All Fields]) OR ("vaginal"[All Fields] AND "cloths"[All Fields]) OR ("menstruation"[Title/Abstract] AND "management"[Title/Abstract]) OR (("menstruation"[Title/Abstract] OR "menstrual"[Title/Abstract]) AND "product*"[Title/Abstract] AND "safety"[Title/Abstract]) OR "catamenia"[All Fields]) AND ("HIV"[MeSH] OR "hiv infections"[MeSH] OR "HIV"[All Fields] OR "Human Immunodeficiency Virus"[All Fields] OR "hiv infections"[All Fields]) |
| Human Papilloma Virus | ("Menstrual Hygiene Products"[MeSH] OR "period product*"[Title/Abstract] OR "feminine hygiene product*"[Title/Abstract] OR "feminine hygiene"[Title/Abstract] OR "feminine product*"[Title/Abstract] OR "feminine napkin*"[Title/Abstract] OR "sanitary pad*"[Title/Abstract] OR "menstrual cup"[Title/Abstract] OR "vaginal cup"[Title/Abstract] OR "menstrual cups"[Title/Abstract] OR "menses cups"[Title/Abstract] OR "vaginal cups"[Title/Abstract] OR "reusable pad*"[Title/Abstract] OR ("menstrual"[All Fields] AND "cloths"[All Fields]) OR ("menses"[All Fields] AND "cloths"[All Fields]) OR ("menstruation"[All Fields] AND "cloths"[All Fields]) OR ("vaginal"[All Fields] AND "cloths"[All Fields]) OR ("menstruation"[Title/Abstract] AND "management"[Title/Abstract]) OR (("menstruation"[Title/Abstract] OR "menstrual"[Title/Abstract]) AND "product*"[Title/Abstract] AND "safety"[Title/Abstract]) OR "catamenia"[All Fields]) AND ("Human Papillomavirus Viruses"[MeSH] OR "Human Papillomavirus"[Title/Abstract] OR "Human Papillomavirus Virus"[Title/Abstract] OR "HPV"[Title/Abstract]) |
| All terms | ( "Menstrual Hygiene Products"[Mesh] OR "menstrual hygiene"[tiab] OR "menstrual product*"[tiab] OR "menstrual materials"[tiab] OR "period product*"[tiab] OR "feminine product*"[tiab] OR "feminine hygiene product*"[tiab] OR "feminine napkin*"[tiab] OR "sanitary pad*"[tiab] OR "sanitary napkin*"[tiab] OR "vaginal cup"[tiab:~1] OR "vaginal cups"[tiab:~1] OR ((menstrual[tiab] OR menses[tiab] OR menstruation[tiab] OR catamenia[tiab] OR "feminine hygiene"[tiab]) AND (cup[tiab] OR cups[tiab] OR cloth*[tiab] OR pad[tiab] OR pads[tiab])) )  AND  ( "HIV"[MeSH] OR "HIV Infections"[MeSH] OR "HIV"[tiab] OR "human immunodeficiency virus*" OR "Syphilis"[Mesh] OR syphilis[tiab] OR "Gonorrhea"[Mesh] OR "Neisseria gonorrhoeae"[Mesh] OR gonorrhea[tiab] OR gonococcus[tiab] OR gonorrhoeae[tiab] OR "Chlamydia"[Mesh] OR chlamydia[tiab] OR "Urinary Tract Infections"[Mesh] OR "urinary tract infection*"[tiab] OR "urogenital infection*"[tiab] OR bacteriuria*[tiab] OR pyuria[tiab] OR "Vaginosis, Bacterial"[MeSH] OR "bacterial vaginosis"[tiab] OR "BV"[tiab] OR "Human Papillomavirus Viruses"[Mesh] OR papilloma*[tiab] OR HPV[tiab] ) |

Supplementary Table 2. Associations between menstrual health products and infectious disease outcomes, stratified by outcome of interest.

| **Outcome^1^** | **Study** | **Comparison Groups** | **Effect Measures^2^** |
| --- | --- | --- | --- |
| **Self-reported symptoms of RTI/STI/UTI** | | | |
|  | Balamurugan 2012 | Difference in prevalence of RTI symptoms, comparing women using cloth vs. sanitary pads | 38% vs. 15%; p-value < 0.05 |
|  | Philip 2013 | 1.) Difference in prevalence of symptoms of RTIs/STIs, comparing women using ordinary cloth vs. sanitary pads  2.) OR of RTI/STI symptoms, comparing ordinary cloth vs. sanitary pads (referent group) | 1.) 20.7% vs. 9.9%; p-value = 0.033  2.) OR: 2.38 (95% CI: 1.00‚ 5.86) |
|  | Das 2015 | aOR of urogenital symptoms, comparing reusable cloths vs. disposable pads (referent group) | aOR = 2.26 (95% CI: 1.50, 3.40) |
|  | Tchoudomirova 1998 | Difference in prevalence of 1.) tampon only use or 2.) sanitary napkin only use, comparing women with vs. without symptoms;  3.) aOR of recurrent urinary symptom, comparing tampon use vs. no tampon use (referent group) | 1.) 16.6% vs. 23.5%; p-value = 0.03  2.) 22.1% vs. 22.0%; p-value = 0.96  3.) aOR = 0.67 (95% CI: 0.44, 1.0) |
|  | Nabwera 2021 | 1.) aPR of UTI symptoms in preceding 24 hours, comparing provision of menstrual pads at school vs no provision of menstrual pads at school (referent group) among adolescent girls  2.) aPR of RTI symptoms in preceding 2 months, comparing provision of menstrual pads at school vs no provision of menstrual pads at school (referent group) among adolescent girls  3.) aPR of UTI symptoms in preceding 24 hours, comparing use of other sanitary material other than disposable pads vs. disposable pads (referent group)  4.) aPR of RTI symptoms in preceding 2 months, comparing use of other sanitary material other than disposable pads vs. disposable pads (referent group) | 1.) aPR = 1.40 (95% CI: 1.20, 1.50); p-value < 0.01  2.) aPR = 1.00 (95% CI: 0.80, 1.30); p-value = 0.76  3.) aPR = 0.90 (95% CI: 0.80, 1.10); p-value = 0.40  4.) aPR = 1.00 (95% CI: 0.80, 1.10); p-value = 0.23 |
|  | Howard 2011 | Difference in prevalence of urovaginal symptoms, comparing women using tampons vs. menstrual cups | 27% vs. 51%; p-value = 0.02 |
|  | Chakrabarty 2023 | Difference in ATT of RTI symptoms, comparing women using hygienic menstrual materials (sanitary napkins, locally made napkins, tampons, and menstrual cups) vs. unhygienic materials (cloth, other materials not included in the hygienic category, or no product use) | 0.032 vs. 0.0416; p-value < 0.05 |
|  | Al Karmi 2024 | OR of UTI/RTI symptoms, comparing reuse of menstrual products vs. no reuse of menstrual products (referent group) | OR = 2.45 (95% CI: 1.50, 4.02);  p-value < 0.01 |
|  | Singh, A 2022 | Difference in prevalence of vaginal discharge, comparing women using only sanitary napkins vs. any other menstrual product | 43.9% vs. 56.1%; p < 0.001 |
|  | Singh, M 2022 | 1.) aOR of having at least one RTI symptoms, comparing women using cloth only vs pads only (referent group)  2.) aOR of having at least one RTI symptoms, comparing women using cloth and pads vs. pads only (referent group) | 1.) aOR = 1.97 (95% CI: 0.77, 5.05); p-value = 0.16  2.) aOR = 1.37 (95% CI: 0.46, 4.04); p-value = 0.57 |
|  | Janoowalla 2020 | 1.) aOR of UTI symptoms without confirmation by urine culture, comparing single-use menstrual pads vs. no menstrual pad use (referent group)  2.) aOR of vulvovaginal symptoms, comparing single-use menstrual pads vs. no menstrual pad use (referent group) | 1.) aOR = 1.02 (95% CI: 0.66, 1.58); p-value = 0.93  2.) aOR = 0.89 (95% CI: 0.52, 1.52); p-value = 0.67 |
| **Confirmed Bacterial Vaginosis** | | | |
|  | Baisley 2009 | 1.) aOR of BV, comparing women using cloths, underwear, or sponges vs. sanitary pads (referent group)  2.) aOR of BV, comparing women using cotton wool or toilet paper vs. sanitary pads (referent group) | 1.) aOR = 1.42 (95% CI: 1.02, 1.95)  2.) aOR = 2.52 (95% CI: 1.21, 5.25) |
|  | Klebanoff 2010 | 1.) aPR of BV, comparing women using tampons only vs. pads only (referent group)  2.) aPR of BV, comparing women using tampons and pads vs. pads only (referent group)  3.) PR of BV, comparing women using pads only vs. women experiencing amenorrhea (referent group)  4.) PR of BV, comparing women using tampons only vs. women experiencing amenorrhea (referent group)  5.) PR of BV, comparing women using tampons and pads vs. women experiencing amenorrhea (referent group) | 1.) aPR = 1.04 (95% CI: 0.95, 1.12)  2.) aPR = 1.00 (95% CI: 0.92, 1.07)  3.) PR = 1.25 (95% CI: 1.16, 1.34)  4.) PR = 1.21 (95% CI: 1.10, 1.33)  5.) PR = 1.13 (95% CI: 1.03, 1.23) |
|  | Torondel 2018 | 1.) Difference in prevalence of BV infection, comparing women using old cotton fabric vs. old silk/nylon fabric as a reusable material  2.) aPRR of BV infection, comparing women using reusable cloths vs. disposable sanitary pads (referent group) | 1.) 42.1% vs. 50.4%; p-value = 0.16  2.) aPRR = 1.23 (95% CI: 1.00, 1.54) |
|  | Das 2021 | 1.) OR of BV infection, comparing women using old silk/nylon (sari or other) vs. old cotton (sari or other; referent group) as reusable product materials  2.) OR of BV infection, comparing women using towels vs. old cotton (sari or other; referent group) as reusable product materials | 1.) OR = 1.3 (95% CI: 0.9‚ 1.8)  2.) OR = 1.0 (95% CI: 0.4, 2.3) |
|  | Das 2015 | aOR of lab confirmed BV, comparing women using reusable cloths vs. disposable pads (referent group) | aOR = 1.23 (95% CI: 0.8, 2.0) |
|  | Demba 2005 | Difference in prevalence of BV, comparing women using sanitary pads vs. traditional methods (old cloths washed and re-used as necessary) | 43.0% vs. 50.7%; p-value = 0.32 |
|  | Morison 2005 | aOR of lab confirmed BV, comparing women using modern sanitary pads vs. traditional cloths (referent group) | aOR = 1.44 (95% CI: 0.97, 2.12); p-value = 0.07 |
|  | Klatt 2010 | OR of recurrent BV, comparing women using menstrual pads vs. no menstrual pad use (referent group) | OR = 8.36 (95% CI: 1.85, 76.34); Association in multivariate model was non-significant |
|  | Phillips-Howard 2016 | 1.) aPR of composite BV at end line, comparing girls receiving insertable menstrual cups vs. monthly sanitary pads (referent group)  2.) aPR of composite BV at end line, comparing girls receiving insertable menstrual cups vs. usual practice (referent group)  3.) aPR of composite BV at end line, comparing girls receiving monthly sanitary pads vs. usual practice (referent group)  4.) aPR of composite BV at end line, comparing girls receiving either insertable menstrual cups or monthly sanitary pads vs. usual practice (referent group) | 1.) aPR = 0.71(95% CI: 0.46, 1.10); p-value = 0.13  2.) aPR = 0.71 (95% CI: 0.46, 1.11); p-value = 0.14  3.) aPR = 1.00 (95% CI: 0.66, 1.53); p-value = 0.99  4.) aPR = 0.85 (95% CI: 0.58, 1.23); p-value = 0.38 |
|  | Mehta 2021 | 1.) Difference in prevalence of pad use during last period, comparing women living with BV vs. women living without BV  2.) Difference in prevalence of cloth use during part or all of last period, comparing women living with BV vs. women living without BV  3.) PRR of BV, comparing women who used cloth to manage last menstrual period vs women who did not use cloth (referent group) | 1.) 95.8% vs. 93.7%; p-value = 0.76  2.) 29.2% vs. 24.5%; p-value = 0.49  3.) PRR = 1.21 (95% CI: 0.78, 1.86) |
|  | Mehta 2023 | aOR of BV, comparing adolescent girls using menstrual cups vs. usual practice (referent group) | aOR = 0.82 (95% CI: 0.51, 1.32); p-value = 0.42 |
| **Confirmed HIV** | | | |
|  | Demba 2005 | Difference in prevalence of HIV, comparing women using sanitary pads vs. traditional methods (old cloths washed and re-used as necessary) | 12.7% vs. 13.2%; p-value = 1.00 |
|  | Phillips-Howard 2015 | Difference in prevalence of menstrual item use (commercial pads vs. traditional pads, including cloth, tissues, paper, locally made or other makeshift items), comparing 1.) women living with HIV vs. 2.) women living without HIV | 1.) 73.2% vs. 26.8%  2.) 60.9% vs. 39.1%  p-value < 0.01 |
|  | Zulaika 2023 | aRR of incident HIV, comparing adolescent girls using 1.) menstrual cups vs. referent group, 2.) CCTs vs referent group, and 3.) combined menstrual cups and CCTs vs referent group, where the referent group were those in the usual practice arm receiving soap for handwashing | 1.) aRR = 0.88 (95% CI: 0.38, 2.05); p-value = 0.77  2.) aRR = 1.16 (95% CI: 0.51, 2.62); p-value = 0.72  3.) aRR = 0.80 (95% CI: 0.33, 1.94); p-value = 0.62 |
| **Self-reported composite UTIs** | | | |
|  | Omar 1998 | 1.) Difference in incidence of UTIs from the time of menarche until the completion of the study survey, comparing women using pads vs. tampons or combination use  2.) Difference in incidence of recurrent UTIs from the time of menarche until the completion of the study survey, comparing women using pads vs. tampons or combination use | 1.) 12.5% vs. 32.2%; p-value < 0.01  2.) 2.1% vs. 13.9%; p-value = 0.02 |
| ***Candida albicans*** | | | |
|  | Phillips-Howard 2016 | 1.) aPR of composite CA at end line, comparing girls receiving insertable menstrual cups vs. monthly sanitary pads (referent group)  2.) aPR of composite CA at end line, comparing girls receiving insertable menstrual cups vs. usual practice (referent group)  3.) aPR of composite CA at end line, comparing girls receiving monthly sanitary pads vs. usual practice (referent group)  4.) aPR of composite CA at end line, comparing girls receiving either insertable menstrual cups or monthly sanitary pads vs. usual practice (referent group) | 1.) aPR = 0.83 (95% CI: 0.31, 2.23); p-value = 0.71  2.) aPR = 0.93 (95% CI: 0.37, 2.38); p-value = 0.89  3.) aPR = 1.13 (95% CI: 0.62, 2.08); p-value = 0.68  4.) aPR = 1.03 (95% CI: 0.56, 1.90); p-value = 0.93 |
|  | Torondel 2018 | 1.) Difference in prevalence of CA infection, comparing women using old cotton fabric vs. old silk/nylon fabric as a reusable material  2.) aPRR of CA infection, comparing women using reusable cloths vs. disposable sanitary pads (referent group) | 1.) 42.8% vs. 42.3%; p-value = 0.94  2.) aPRR = 1.54 (95% CI: 1.21, 2.00) |
| **Confirmed composite STIs** | | | |
|  | Phillips-Howard 2016 | 1.) aPR of composite STIs at end line, comparing girls receiving insertable menstrual cups vs. monthly sanitary pads (referent group)  2.) aPR of composite STIs at end line, comparing girls receiving insertable menstrual cups vs. usual practice (referent group)  3.) aPR of composite STIs at end line, comparing girls receiving monthly sanitary pads vs. usual practice (referent group)  4.) aPR of composite STIs at end line, comparing girls receiving either insertable menstrual cups or monthly sanitary pads vs. usual practice (referent group) | 1.) aPR = 0.78 (95% CI: 0.36, 1.68);  p-value = 0.52  2.) aPR = 0.48 (95% CI: 0.24, 0.96); p-value = 0.04  3.) aPR = 0.62 (95% CI: 0.37, 1.03); p-value = 0.06  4.) aPR = 0.54 (95% CI: 0.34, 0.87); p-value = 0.01 |
|  | Mehta 2021 | 1.) Difference in prevalence of pad use during last period, comparing women diagnosed with an STI vs. women not diagnosed with an STI  2.) Difference in prevalence of cloth use during part or all of last period, comparing women diagnosed with an STI vs. women not diagnosed with an STI | 1.) 95.1% vs. 93.9%; p-value > 0.99  2.) 31.7% vs. 24.4%; p-value = 0.30 |
|  | Mehta 2023 | aRR of composite STIs, comparing adolescent girls using menstrual cups vs. usual practice (referent group) | aRR = 0.77 (95% CI: 0.62, 0.95) |
| ***Chlamydia trachomatis*** | | | |
|  | Phillips-Howard 2016 | 1.) aPR of composite CT at end line, comparing girls receiving insertable menstrual cups vs. monthly sanitary pads (referent group)  2.) aPR of composite CT at end line, comparing girls receiving insertable menstrual cups vs. usual practice (referent group)  3.) aPR of composite CT at end line, comparing girls receiving monthly sanitary pads vs. usual practice (referent group)  4.) aPR of composite CT at end line, comparing girls receiving either insertable menstrual cups or monthly sanitary pads vs. usual practice (referent group) | 1.) aPR = 1.15 (95% CI: 0.29, 4.57); p-value = 0.85  2.) aPR = 0.39 (95% CI: 0.12, 1.23); p-value = 0.11  3.) aPR = 0.34 (95% CI: 0.12, 0.96); p-value = 0.04  4.) aPR = 0.36 (95% CI: 0.15, 0.86); p-value = 0.02 |
| **Changes in Vaginal Microbiome** | | | |
|  | Unzeitig 2007 | 1.) Difference in microbial colonization of the vagina, taken just before the onset of menstruation, comparing women using sanitary towels vs. tampons only  2.) Difference in microbial colonization of the vagina, taken during menstruation, comparing women using sanitary towels vs. tampons only | 1.) Colonization was entirely comparable between groups, except for the presence of *Escherichia coli* (20% vs. 16%) and *enterococci* (10% vs. 7%)  1.) Colonization was entirely comparable between groups, except for slightly lower occurrence of *peptostreptococci*, a significantly lower occurrence of *Escherichia coli* (34% vs. 18%) and *enterococci* (24% vs. 9%) |
|  | Mehta 2023 | 1.) OR of CST-I vs. other CST, comparing adolescent girls using menstrual cups vs. usual practice (referent group)  2.) Mean relative abundance of *Lactobacillus crispatus*, comparing adolescent girls using menstrual cups vs. usual practice (referent group) | 1.) aOR = 1.42 (95% CI: 1.21, 1.67)  2.) Mean Relative Abundance = 4.46 (95% CI: 2.76, 6.16) |
| **Confirmed UTIs and/or BV** | | | |
|  | Das 2015 | aOR of lab confirmed BV and/or UTIs, comparing reusable cloths vs. disposable pads (referent group) | aOR = 2.8 (95% CI: 1.7, 4.5) |
| **Human Papillomavirus** | | | |
|  | Abulizi 2017 | aOR of HPV infection, comparing women using sanitary napkins or toilet paper vs. cloth (referent group) | aOR = 0.58 (95% CI: 0.37, 0.93) |
| ***Gardnerella vaginalis*** | | | |
|  | Hansen 1985 | Difference in GV prevalence, comparing women using tampons vs. other types of menstrual hygiene products | 61.9% vs. 48.8%; p-value < 0.01 |
| ***coag-neg Staph*** | | | |
|  | Leibovici 1984 | Difference in prevalence of tampon use, comparing women with coag-neg Staph vs. women with other causes of UTI (referent group) | 47.8% vs. 80.0%; p-value = 0.03 |
| **Confirmed composite UTIs** | | | |
|  | Foxman 1995 | 1.) OR of first-time UTI, comparing women using tampons only vs. sanitary napkins only (referent group)  2.) OR of first-time UTI, comparing women using sanitary napkins and tampons vs. sanitary napkins only (referent group)  3.) OR of first-time UTI, comparing women using deodorant napkins or tampons vs. sanitary napkins only (referent group) | 1.) OR = 0.57 (95% CI: 0.22, 1.49)  2.) OR = 0.57 (95% CI: 0.25, 1.28)  3.) OR = 1.51 (95% CI: 0.74, 3.06) |
|  | Das 2015 | aOR of lab confirmed UTIs, comparing reusable cloths vs. disposable pads (referent group) | aOR = 2.0 (95% CI: 1.0, 4.0) |
|  | Janoowalla 2020 | aOR of UTI (positive urine culture), comparing single-use menstrual pads vs. no menstrual pad use (referent group) | aOR = 2.09 (95% CI: 0.89, 4.91);  p-value = 0.09 |
|  | Madar 2024 | Difference in prevalence of UTIs, comparing women using menstrual cups vs. non-users/not regular users (referent group) | 36.0% vs. 34.6%; p-value = 0.90 |
| **Confirmed composite RTIs** | | | |
|  | Phillips-Howard 2016 | 1.) aPR of composite RTIs at end line, comparing girls receiving insertable menstrual cups vs. monthly sanitary pads (referent group)  2.) aPR of composite RTIs at end line, comparing girls receiving insertable menstrual cups vs. usual practice (referent group)  3.) aPR of composite RTIs at end line, comparing girls receiving monthly sanitary pads vs. usual practice (referent group)  4.) aPR of composite RTIs at end line, comparing girls receiving either insertable menstrual cups or monthly sanitary pads vs. usual practice (referent group) | 1.) aPR = 0.73 (95% CI: 0.46, 1.15); p-value = 0.18  2.) aPR = 0.79 (95% CI: 0.48, 1.30); p-value = 0.36  3.) aPR = 1.09 (95% CI: 0.80, 1.48); p-value = 0.58  4.) aPR = 0.93 (95% CI: 0.66, 1.31); p-value = 0.68 |
| **Self-reported composite STIs** | | | |
|  | Omar 1998 | Difference in incidence of STIs from the time of menarche until the completion of the study survey, comparing women using pads vs. tampons or combination use | 12.5% vs. 14.9%; p-value = 0.38 |
|  | Janoowalla 2020 | aOR of self-reported history of STIs, comparing single-use menstrual pads vs. no menstrual pad use (referent group) | aOR = 1.86 (95% CI: 0.67, 5.20);  p-value = 0.24 |
| **Vulvovaginal candidiasis** | | | |
|  | Geiger 1996 | 1.) aOR of any sanitary napkin use during last menses, comparing women with VVC vs. population controls without VVC (referent group)  2.) aOR of baking soda-treated sanitary napkin use during last menses, comparing women with VVC vs. population controls without VVC (referent group)  3.) aOR of deodorant sanitary napkin use during last menses, comparing women with VVC vs. population controls without VVC (referent group)  4.) aOR of any tampon use during last menses, comparing women with VVC vs. population controls without VVC (referent group)  5.) aOR of deodorant tampon use during last menses, comparing women with VVC vs. population controls without VVC (referent group) | 1.) aOR = 1.30 (95% CI: 0.58-2.91)  2.) aOR = 2.16 (95% CI: 0.53-8.82)  3.) aOR = 1.70 (95% CI: 0.78-3.70)  4.) aOR = 1.03 (95% CI: 0.50, 2.12)  5.) aOR = 1.94 (95% CI: 0.84, 4.50) |
|  | Das 2021 | 1.) OR of VVC infection, comparing women using old silk/nylon (sari or other) vs. old cotton (sari or other; referent group) as reusable product materials  2.) OR of VVC infection, comparing women using towels vs. old cotton (sari or other; referent group) as reusable product materials | 1.) OR = 1.0 (95% CI: 0.7, 1.4)  2.) OR = 1.0 (95% CI: 0.5, 2.3) |
| ***Neisseria gonorrhoeae*** | | | |
|  | Phillips-Howard 2016 | 1.) PR of composite GC at end line, comparing girls receiving insertable menstrual cups vs. monthly sanitary pads (referent group)  2.) PR of composite GC at end line, comparing girls receiving insertable menstrual cups vs. usual practice (referent group)  3.) PR of composite GC at end line, comparing girls receiving monthly sanitary pads vs. usual practice (referent group)  4.) PR of composite GC at end line, comparing girls receiving either insertable menstrual cups or monthly sanitary pads vs. usual practice (referent group) | 1.) PR = 1.40 (95% CI: 0.10, 19.57); p-value = 0.81  2.) PR = 1.07 (95% CI: 0.10, 11.56); p-value = 0.96  3.) PR = 0.77 (95% CI: 0.08, 7.24); p-value = 0.82  4.) PR = 0.91 (95% CI: 0.14, 6.06); p-value = 0.92 |
| ***Trichomonas vaginalis*** | | | |
|  | Phillips-Howard 2016 | 1.) PR of composite TV at end line, comparing girls receiving insertable menstrual cups vs. monthly sanitary pads (referent group)  2.) PR of composite TV at end line, comparing girls receiving insertable menstrual cups vs. usual practice (referent group)  3.) PR of composite TV at end line, comparing girls receiving monthly sanitary pads vs. usual practice (referent group)  4.) PR of composite TV at end line, comparing girls receiving either insertable menstrual cups or monthly sanitary pads vs. usual practice (referent group) | 1.) PR = 0.56 (95% CI: 0.16, 1.96); p-value = 0.36  2.) PR = 0.36 (95% CI: 0.11, 1.28); p-value = 0.12  3.) PR = 0.65 (95% CI: 0.21, 2.02); p-value = 0.46  4.) PR = 0.49 (95% CI: 0.18, 1.35); p-value = 0.17 |
|  | Torondel 2018 | 1.) Difference in prevalence of TV infection, comparing women using old cotton fabric vs. old silk/nylon fabric as a reusable material  2.) aPRR of TV infection, comparing women using reusable cloths vs. disposable sanitary pads (referent group) | 1.) 6.2% vs. 7.3%; p-value = 0.72  2.) aPRR = 1.78 (95% CI: 0.81, 3.90) |
| **Herpes Simplex Virus 2** | | | |
|  | Zulaika 2023 | aRR of incident HSV-2, comparing adolescent girls using 1.) menstrual cups vs. referent group, 2.) CCTs vs referent group, and 3.) combined menstrual cups and CCTs vs referent group, where the referent group were those in the usual practice arm receiving soap for handwashing | 1.) aRR = 0.71 (95% CI: 0.50, 1.01); p-value = 0.06  2.) aRR = 1.02 (95% CI: 0.73, 1.41); p-value = 0.92  3.) aRR = 1.16 (95% CI: 0.85, 1.58); p-value = 0.36 |
| ^1^ Confirmed outcome refers to any outcome of interest defined via rapid diagnostic testing, laboratory testing, clinical diagnosis, or other sensitive measurement. Self-reported outcomes refer to any outcomes of interest that were ascertained by participant self-report during study procedures.  ^2^ Adjusted effect measures (aOR, aRR, aPR) are reported, where possible. For studies without adjusted effect measures available for data extraction, crude estimates were included where possible. All effect estimates were rounded to a maximum of two significant figures for consistency.  **Abbreviations:** RTI, reproductive tract infection; STI, sexually transmitted infection; UTI, urinary tract infection; OR, odds ratio; ATT, Average Treatment Effect on the Treated; aOR, adjusted odds ratio; aPR, adjusted prevalence ratio; BV, bacterial vaginosis; PR, prevalence ratio; PRR, prevalence rate ratio; aRR, adjusted relative risk; HIV, human immunodeficiency virus; CCT, conditional cash transfer; CST, vaginal community state types; HPV, human papillomavirus; VVC, vulvovaginal candidiasis; CA, *Candida albicans*, GV, *Gardnerella vaginalis*; coag-neg Staph, coagulase-negative *Staphylococcus*; CT, *Chlamydia trachomatis*; GC, *Neisseria gonorrhoeae*; TV, *Trichomonas vaginalis*; HSV-2, herpes simplex virus 2. | | | |
